# Costing Methods for Artificial Intelligence: Systematic Review and Recommended Cost Inventory for in Health Technology Assessment

**DOI:** 10.1101/2025.10.27.25338867

**Authors:** John Tayu Lee, Toby Kai-Bo Shen, Valerie Tzu-Ning Liu, Tony Hsiu-Hsi Chen, Chien-Chang Lee, Chunling Lu, Chang-Wei Huang, Rifat Atun

## Abstract

Economic evaluations of artificial intelligence (AI) in healthcare are expanding rapidly, yet underlying costing methods remains heterogenous, and frequently incomplete for health technology assessment (HTA) and policy decision-making. In our systematic review of 55 studies published between 2010 and 2025, we found that fewer than half of the studies reported explicit costing methods; most pricing analyses failed to describe the basis of fees, subscription terms, or duration of coverage; and few analyses distinguished between average and incremental costs or accounted for economies of scale. Lifecycle expenditures—including development, validation, integration, maintenance, retraining, and decommissioning—were largely omitted, while electricity consumption, data hosting, and cloud infrastructure costs were almost never considered. Sensitivity analysis was the exception rather than the norm, and reporting of cost offsets such as reduced hospital admissions or workforce time savings was inconsistent.

To address these gaps, we propose a 20-item reporting checklist to standardise the costing and pricing of AI interventions. The checklist complements existing HTA frameworks while capturing features unique to AI, such as continuous retraining, reliance on data infrastructure, and recurrent maintenance. We also introduce an AI Costing Inventory and Calculator that operationalises a lifecycle approach, enabling systematic recording of resource use, unit costs, inflation adjustments, and total and incremental costs, including offsets. These tools extend the emerging CHEERS-AI reporting framework by embedding a lifecycle perspective into costing, thereby enabling consistent estimation of resource and cos components and strengthening the methodological foundations of AI economic evaluation for policy use.

## INTRODUCTION

Artificial intelligence (AI) has become a transformative driver of innovation in modern health care^1 2^, utilizing computational techniques, often rooted in machine learning, to enable machines to perform tasks once dependent on human cognition. Across clinical and operational domains, AI is increasingly deployed to interpret complex data, support diagnostic decisions, inform therapeutic reasoning, and optimize workflow efficiency^3 4^. A growing body of evidence demonstrates its technical feasibility, clinical effectiveness, and capacity for integration across key areas of healthcare^5-7^.

Yet despite this progress, the integration of AI into routine practice remains limited. As AI applications progress toward large-scale implementation and come under the scrutiny of health technology assessment (HTA) frameworks, economic evaluation emerges as a critical determinant of value for money and reimbursement eligibility^8^. For health insurers and public payers, robust cost evidence is indispensable to justify coverage and guide investment decisions^9^. Within this context, the generation of accurate, transparent, and reproducible cost estimates becomes paramount^10^.

Although economic evaluation methods are well established in healthcare, they have traditionally focused on discrete interventions such as pharmaceuticals, medical devices, and surgical procedures^11^. These conventional approaches may be inadequate for AI technologies, which often involve iterative development cycles, integration with existing data systems, and ongoing maintenance or recalibration. Therefore, estimating cost for AI intervention remains a largely unmeasured yet critical factor—often described as the “elephant in the room” of AI implementation^12 13^. Without standardized, transparent, and reproducible costing approaches, health systems risk miscalculating the required investment or overlooking downstream cost offsets.

This study aims to: (1) identify and synthesize how the resource use associated with AI-enabled or assisted healthcare interventions have been calculated and reported in the literature; (2) evaluate the methodological approaches used to estimate or price AI technologies; (3) assess whether existing studies account for key cost components, including development, implementation, maintenance, and algorithm retraining; (4) examine gaps and inconsistencies in current costing approaches through a systematic review of published studies; (5) provide recommendations for standardized reporting criteria and propose cost-collection inventory to guide the economic evaluation of AI interventions.

## METHODS

### Protocol and registration

This systematic review was conducted according to a pre-specified protocol and registered with the International Prospective Register of Systematic Reviews (PROSPERO; registration number CRD420251109715).

### Search strategy

A comprehensive search strategy was designed to identify relevant literature across multiple bibliographic databases, including PubMed, EMBASE, and Scopus, as well as specialist health technology assessment (HTA) repositories such as International HTA Database (INAHTA) and NHS Economic Evaluation Database (NHS EED). The search covered publications from 1 January 2010 to October 2025. Reference lists of included studies were additionally screened to capture potentially eligible articles not retrieved by the database search.

The search strategy combined controlled vocabulary (for example, Medical Subject Headings [MeSH]) with free-text terms. Three domains were specified and intersected: AI technologies (artificial intelligence, machine learning, digital health, natural language processing), economic evaluation (costs, economic evaluation, budget impact, HTA), and healthcare settings (healthcare, medical, public health). Full search strategies for each database are reported in the *Supplementary Table 1*.

### Eligibility criteria

Study eligibility was defined a priori using the PICOS framework. The population included any healthcare setting or patient group where AI interventions had been deployed. The intervention of interest was any AI-based technology applied to diagnosis, treatment, prediction, decision support, or related healthcare functions. Comparators, where reported, included standard care or alternative interventions. Outcomes of interest were economic in nature, focusing specifically on cost components, costing methodologies, pricing strategies, and reported cost summaries. Eligible study designs were peer-reviewed primary research articles reporting formal economic evaluations, including cost-effectiveness, cost–utility, cost–benefit, budget impact, and dedicated costing studies. Studies were included if they reported any costing, pricing, or resource-use information related to AI interventions in healthcare, regardless of whether outcome data were available. Studies reporting full or partial economic evaluations (e.g., cost analysis, cost-effectiveness, cost-utility, cost–benefit, or budget impact) were eligible. Commentaries, reviews, and non-original works were excluded.

### Study selection and data extraction

Titles and abstracts were initially screened by one reviewer (TS), with full-text articles assessed against the inclusion criteria. The final set of included studies was reviewed and discussed by two authors (TS, JTL) to confirm eligibility and resolve any uncertainties.

Study characteristics included author(s), year of publication, country or region, and healthcare setting (e.g. hospital, primary care, community health). Intervention characteristics captured the nature of the AI technology (computer-aided detection, decision support, risk prediction and stratification tools, wearable monitoring, workflow optimization software, robotics), the relevant clinical specialty or disease domain (radiology, oncology, ophthalmology, cardiology, infectious disease), and the target population (children, adults, older adults, or unspecified). Economic evaluation characteristics included the type of analysis conducted (costing analysis, cost-effectiveness, cost–utility, cost-benefit analysis) the analytic perspective adopted (health system, payer, societal, patient, or company).

### Costing and Pricing Approaches

In this analysis, we defined resource use narrowly, focusing only on the incremental costs directly related to the AI system (e.g., acquisition, implementation, and maintenance). Routine provider costs that would occur regardless of AI use were excluded.

At the outset, we distinguished between costing and pricing. Costing was defined as the process of identifying, measuring, and valuing the resources actually used to develop, implement, and maintain an AI intervention, typically from the healthcare provider, and payer. The resource-based approach captures the real inputs required to deliver the intervention. Pricing, by contrast, refers to the mechanisms by which a monetary value is assigned to the AI intervention in a market or policy setting. It reflects the amount charges by the developer or vendor, rather than observed resource use.

For costing, we extracted information on the methodological approach (e.g., micro-costing, gross costing, activity-based costing, time-driven activity-based costing, or life-cycle costing) and the cost components included. Micro-costing involves detailed identification and valuation of each resource used (e.g., staff time, equipment, software licensing), providing high accuracy but requiring intensive data collection. Gross costing (or top-down costing) allocates average costs from aggregated financial data, offering simplicity but lower precision. Activity-based or time-driven approaches bridge these by linking costs to clinical activities or time units, improving traceability while reducing data burden. Lifecycle costing extends this further to capture long-term expenses such as retraining, maintenance, and decommissioning.

For pricing analyses, we recorded whether pricing information was reported and, where available, the strategies employed — including subscription-based models, negotiated payment arrangements with public or private payers, benchmarking against existing comparators, and alternative pricing mechanisms. We also captured the level of reporting (e.g., per patient, per use, institutional license, or national procurement) and the underlying basis for price determination, categorized as fixed, negotiated, bundled, or outcome-linked. Where specified, we documented whether the reported prices reflected pre- or post-reimbursement levels or incorporated subsidy arrangements. In addition, we extracted information on whether studies incorporated cost offsets, such as, reduced diagnostic testing, or workforce time savings, and whether sensitivity analyses were conducted to assess parameter uncertainty and robustness of results.

## RESUTLS

We retrieved 11927 citations from bibliographic databases. After removing duplicates, we screened 9005 unique citations by title and abstract, resulting in 624 full-text articles for further evaluation. Out of these, 579 studies were excluded based on predefined criteria, and ultimately, 55 studies met the final inclusion criteria. The PRISMA flowchart of the study identification process is presented in *Fig. 1*.

**Figure 1:**
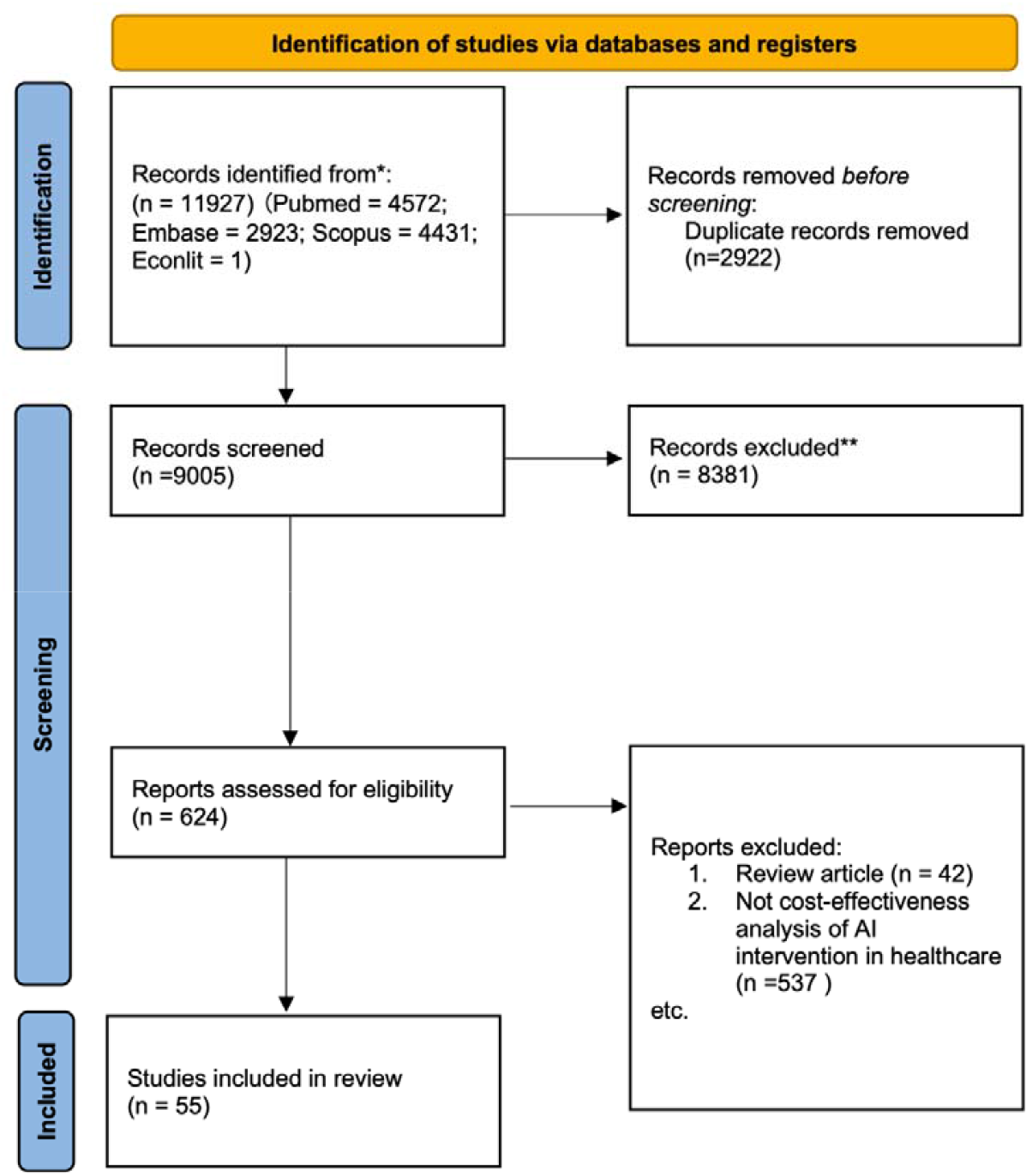
PRISMA Flowchart for Systematic Review

### Characteristics of included articles

The majority of studies evaluated computer-aided detection systems (n = 18)^14-31^, followed by diagnostic support tools (n = 22)^32-53^ and risk prediction or stratification software (n = 9)^54-62^. Less frequently studied interventions included administrative and workflow software (n = 0), robotics (n = 1)^63^, and patient monitoring devices or wearables (n = 1)^64^. Radiology was the predominant clinical specialty (n = 16)^14 15 19 21 24 25 27-29 33 36 37 42 44 50 54 59^, followed by oncology (n = 2)^17 49^, ophthalmology (n = 18)^18 20 22 26 30 34 35 39-41 43 47 48 51 52 61 62 65^, cardiology (n = 3)^46 57 58^, internal medicine (n = 4)^31 32 42 45 53 56^, and infectious disease (n = 4)^16 23 60 64^. Three studies focused on other specialties outside these categories^34 55 63^.

With respect to target populations, most studies assessed adults (n = 45)^14 15 18-29 31 32 35-38 42-51 53 55-60 62 64^, while Seven focused on older populations^17 33 39-41 61 63^ and two on children and adolescents^30 52^. Two studies did not specify the population group^16 54^. The summarize of study characteristics is presented in *Supplementary Table S2*.

Regarding evaluation type, cost–utility analysis was the most frequently applied (n = 13)^17 18 20-22 26 33 38-41 58 60^, followed by cost-effectiveness analysis (n = 31)^14 15 19 23-25 27-32 34-36 43-51 53 55-57 59 63 64^, costing and cost-minimization analyses (n = 5)^16 37 52 61 62^. Few studies assumed no incremental cost for the AI component, typically when the algorithm was embedded within existing diagnostic technologies^31 54 17 20-22 25 26 29 33 36 37 45 46 49 52 53 55 57 58^.

### Costing Methods and Components Included

Approaches to costing varied across studies (*Table 2*). Micro-costing was most common (n = 11)^16 19 30 35 38 39 47 48 50 51 62 63^, followed by gross or top-down methods (n = 8)^18 28 40 41 43 44 61 66 67^, with one study^39^ using an activity-based framework.

**Table 1:** Characteristics of Included Studies.

| Types of AI Intervention | Number of Studies |
| --- | --- |
| Computer Aided Detection (CAD) | 18 |
| Diagnosis Supporting Tool | 22 |
| Risk Prediction and Stratification Software | 9 |
| Patient Monitoring and Wearable | 1 |
| Administrative and Workflow Software | 0 |
| Robotics | 1 |
| **Clinical Area** |  |
| Radiology and Oncology | 18 |
| Internal Medicine | 6 |
| Ophthalmology | 18 |
| Cardiology | 3 |
| Infectious Disease | 4 |
| Mental Health | 2 |
| Others | 1 |
| **Publication date** |  |
| 2020-2025 | 45 |
| 2015-2019 | 3 |
| 2010-2014 | 2 |
| 2005-2009 | 0 |
| 2000-2004 | 0 |
| **Country** |  |
| USA | 15 |
| China | 10 |
| India | 0 |
| UK | 7 |
| European Countries | 6 |
| Canada | 1 |
| Japan | 4 |
| Others | 7 |
| **Targeted Population** |  |
| Not specific | 2 |
| Children and adolescents | 2 |
| Adults | 39 |
| Older Population | 7 |
| **Perspectives** |  |
| Healthcare Payer | 32 |
| Societal | 16 |
| Patients | 1 |
| Companies/Provider/Industry | 0 |
| **Types of Economic Evaluation** |  |
| Costing or Cost-Minimization Analysis | 5 |
| Cost-Consequence Analysis | 1 |
| Cost-Effectiveness Analysis | 31 |
| Cost-Utility Analysis | 13 |
| **AI Intervention Valuation** |  |
| **Approaches** |  |
| Not reported | 10 |
| Not clear | 11 |
| Mainly Costing | 21 |
| Mainly Pricing | 11 |

**Table 2:**
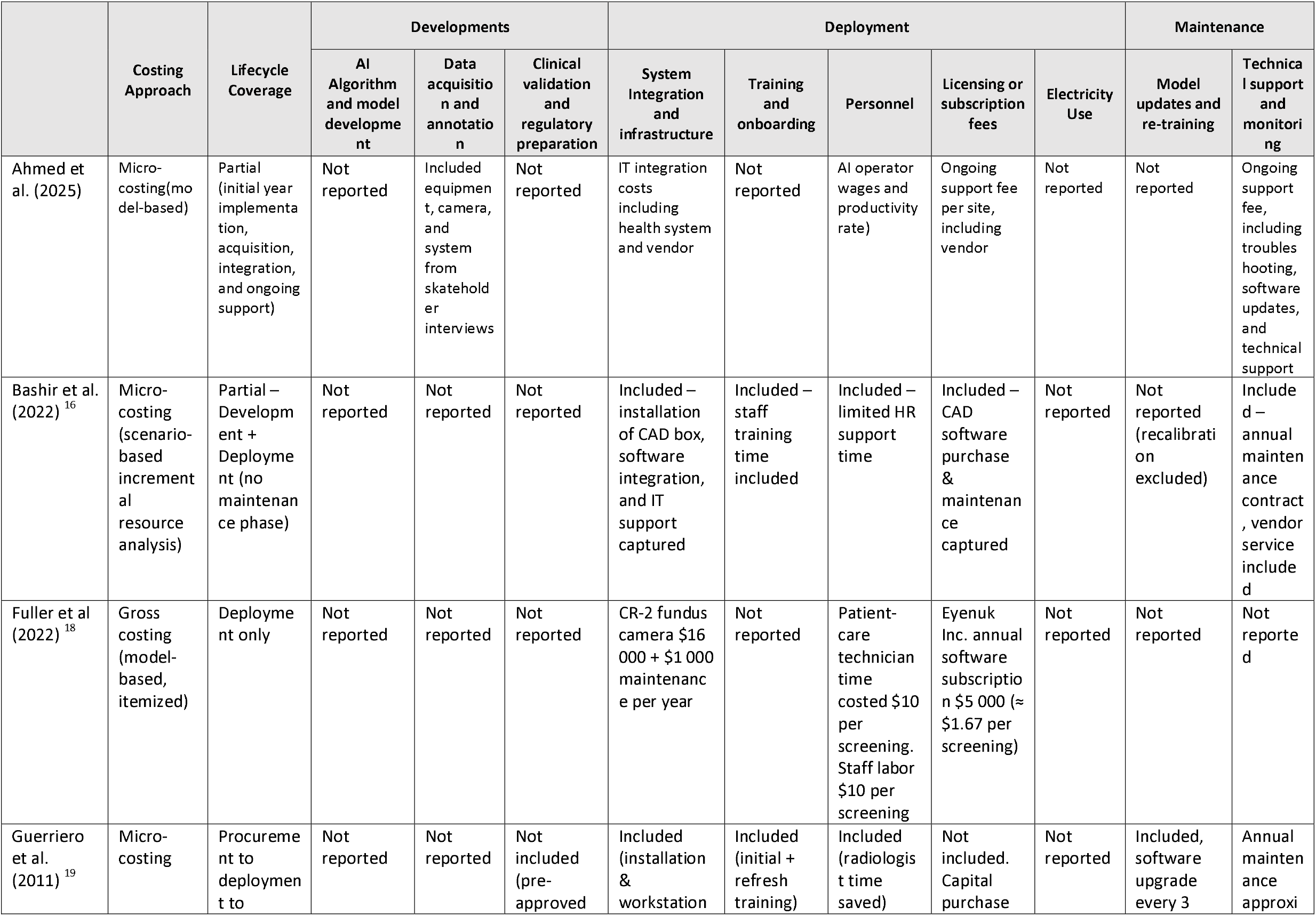
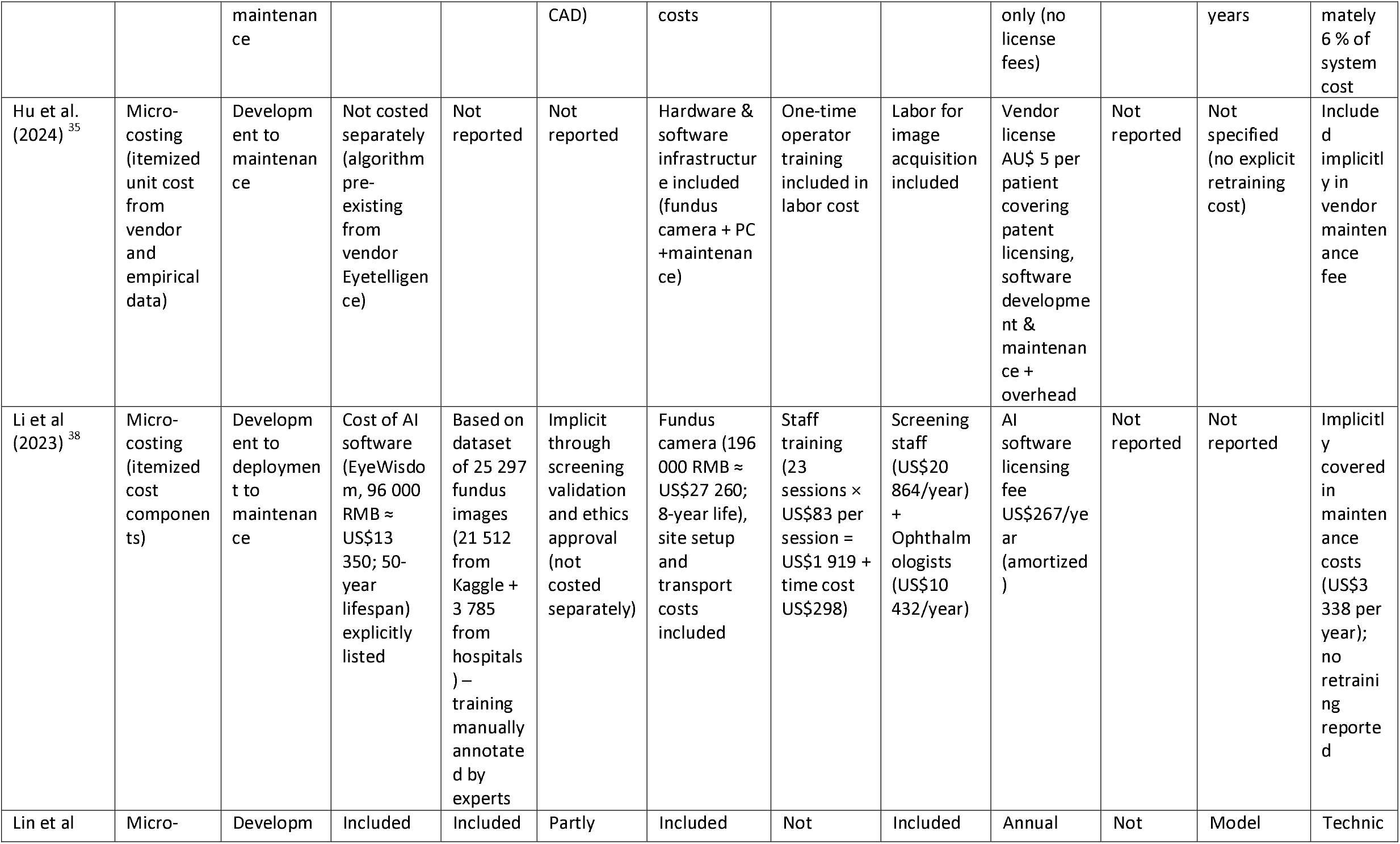
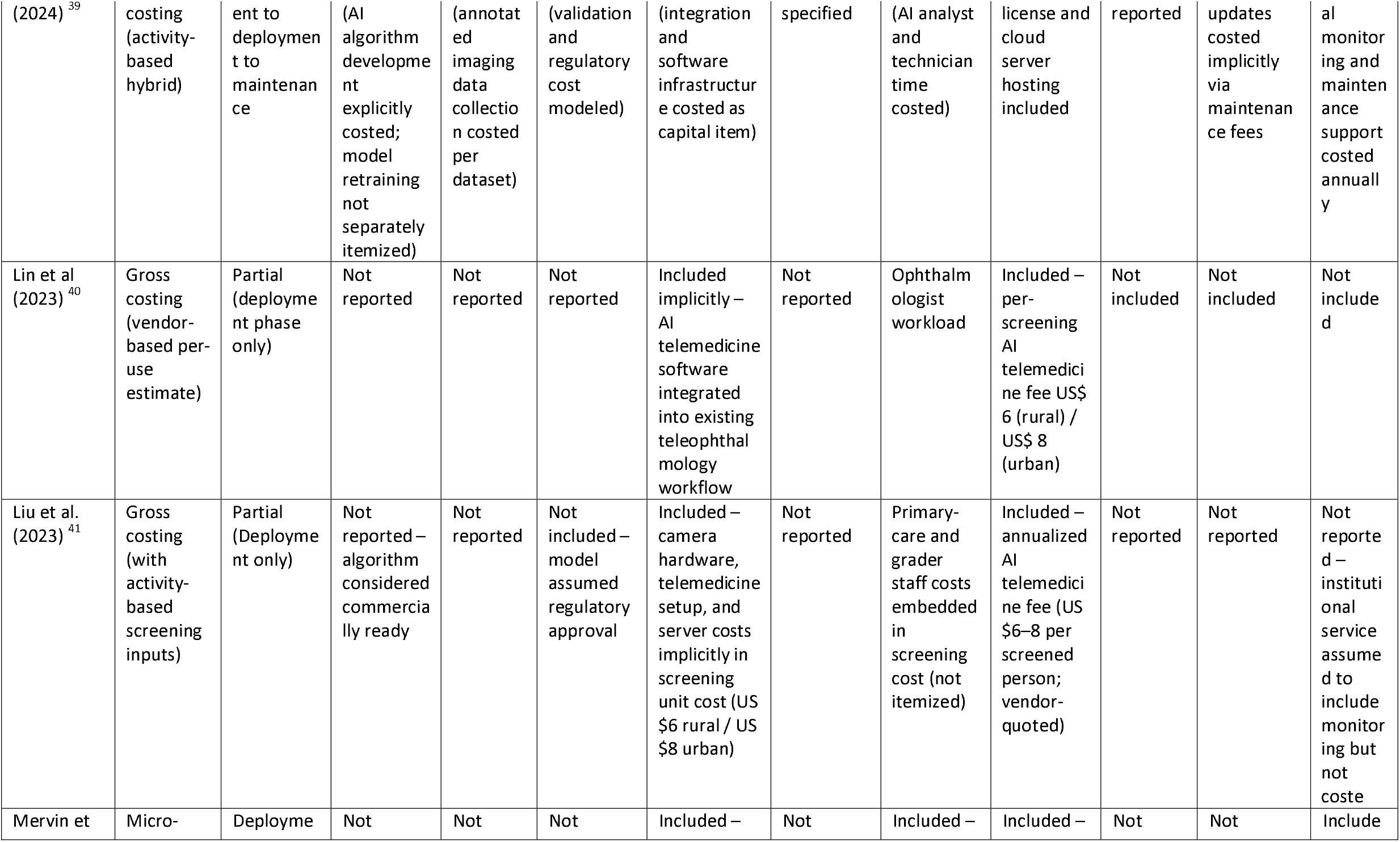
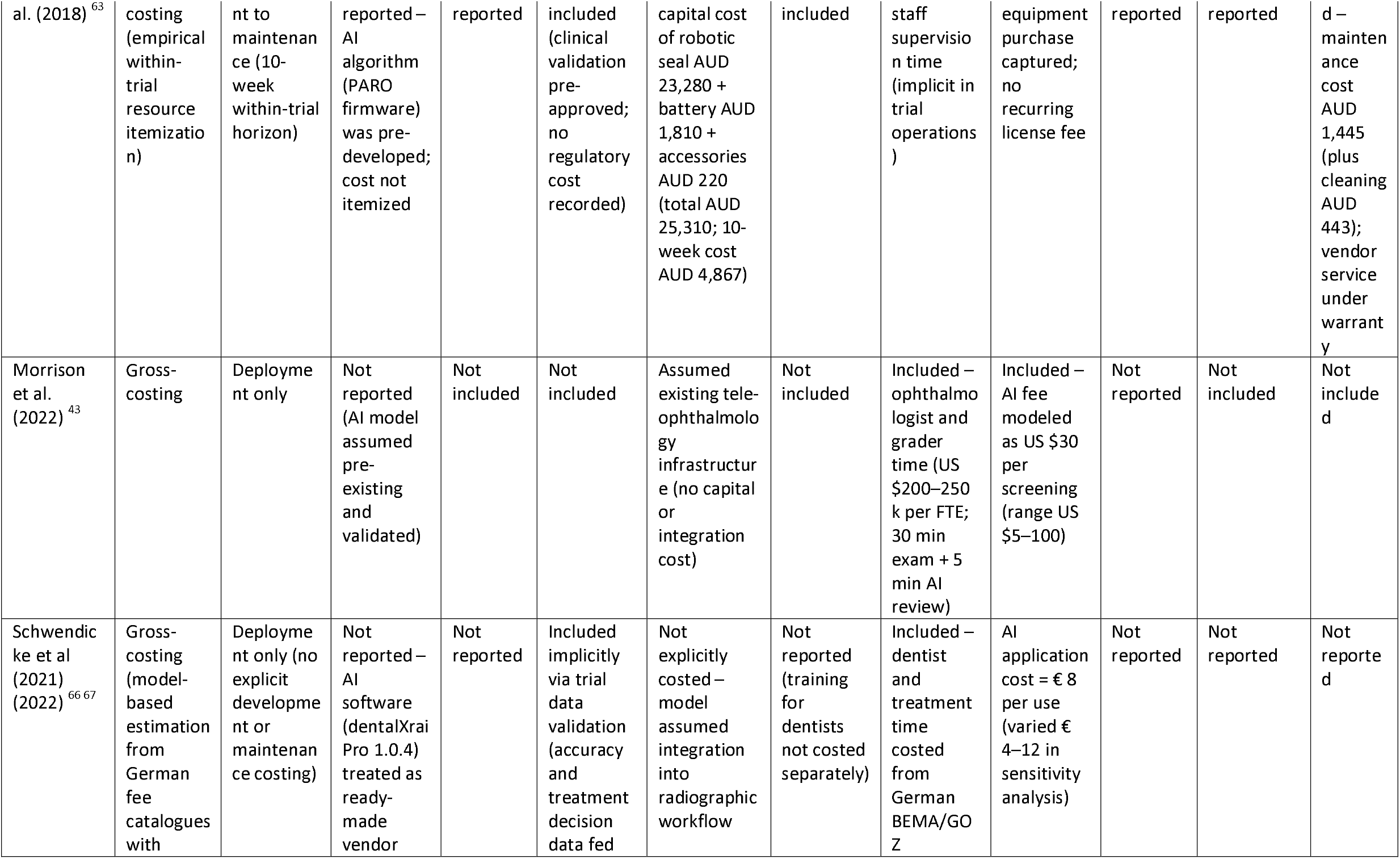
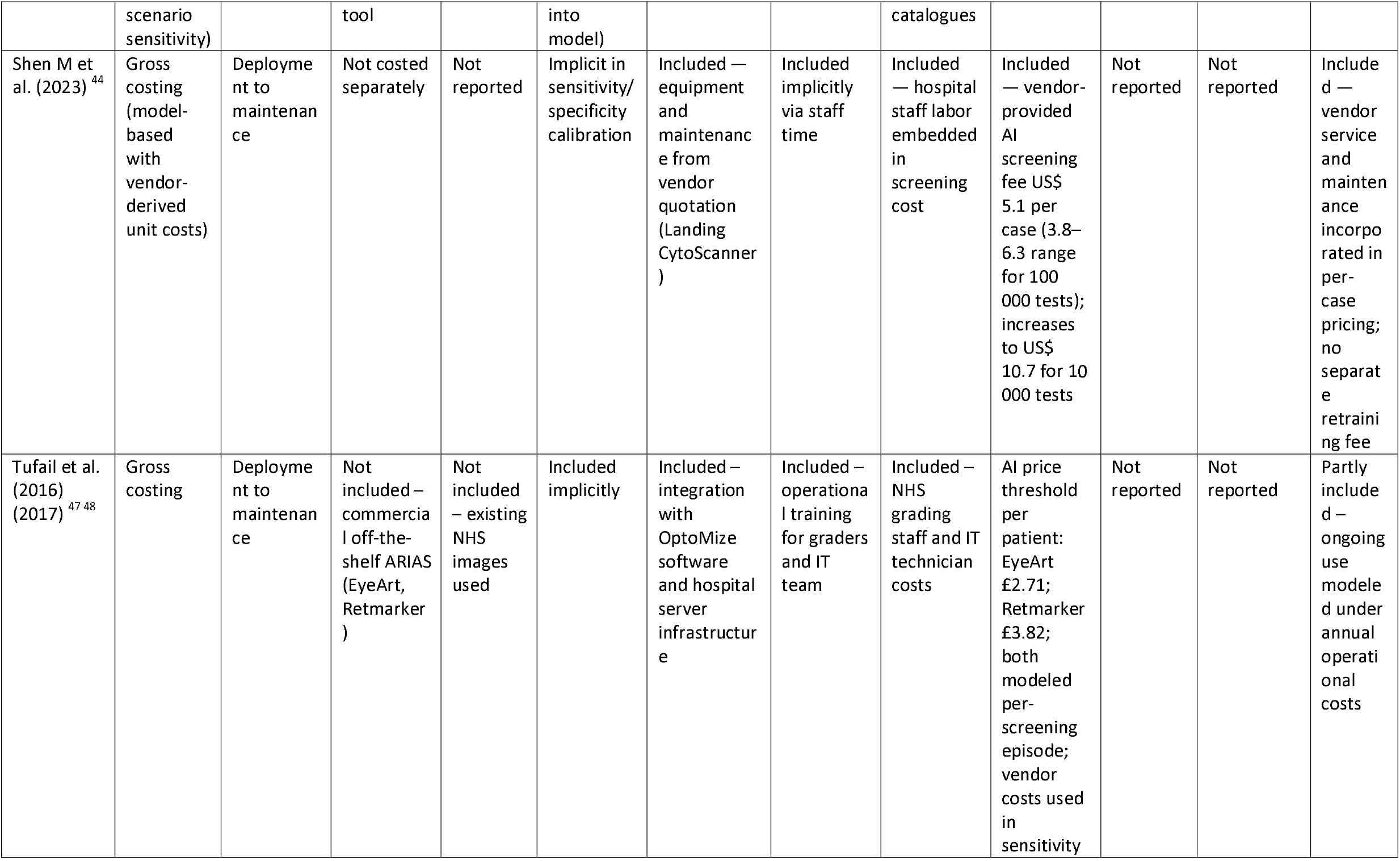
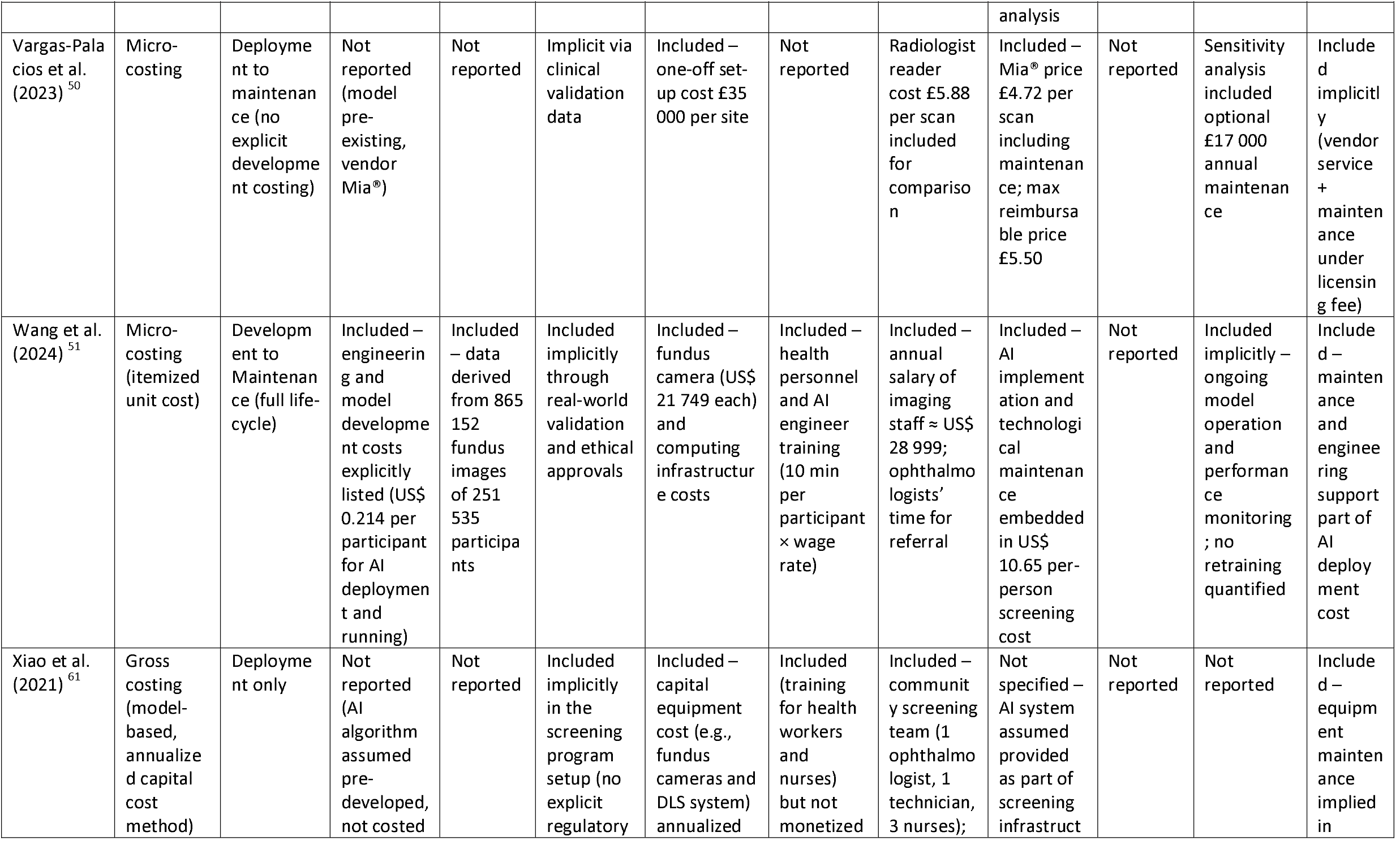
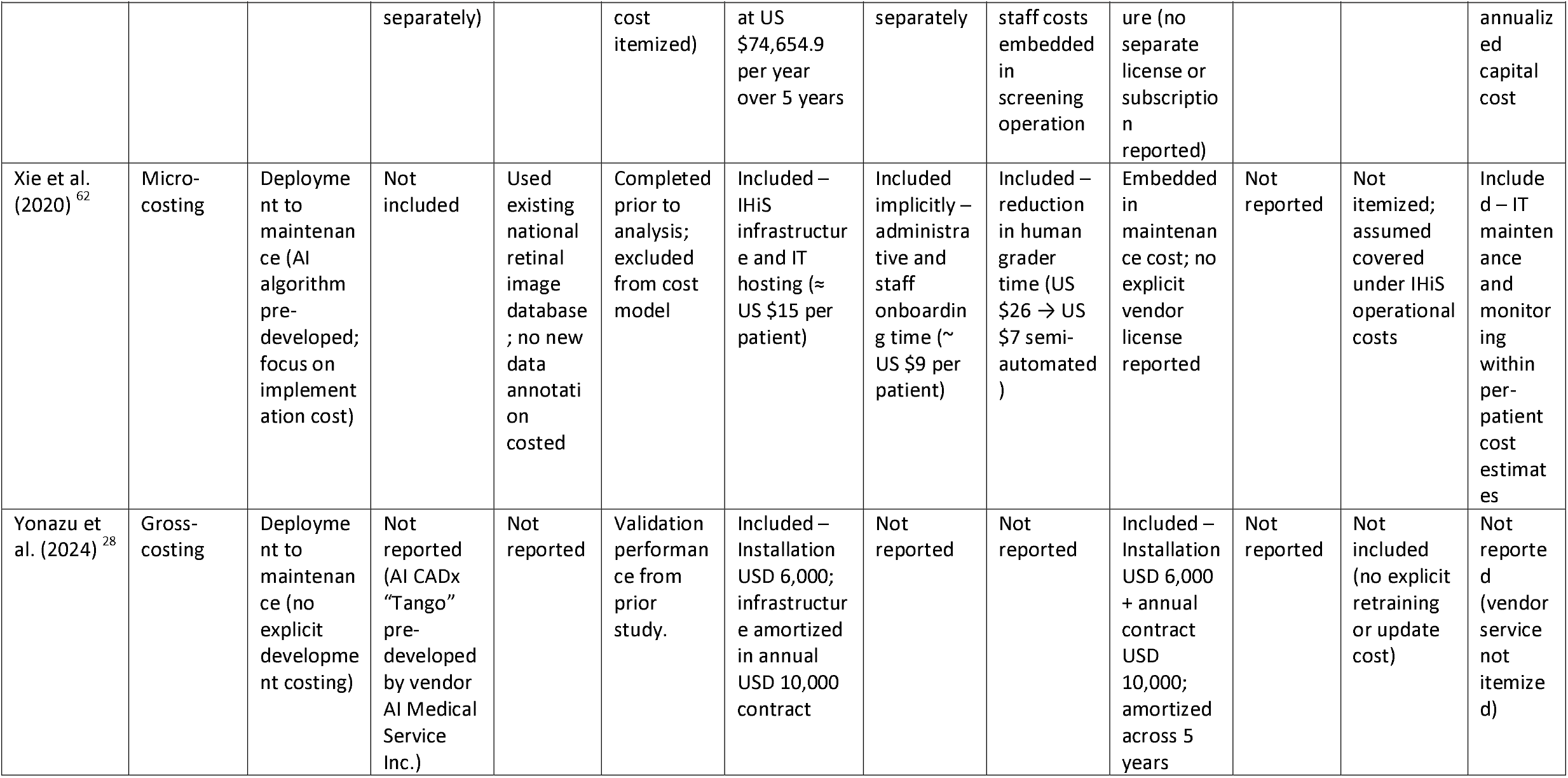
Costing Methods and Components Reported.

Among studies adopting micro-costing, several offered detailed and transparent breakdowns of cost elements. Schwendicke et al. (2022)^66^ itemized development costs—including data labeling, model training, software engineering, and regulatory preparation—alongside cloud infrastructure and support. These were amortized across 100,000 use cases and combined with overheads, yielding a per-analysis cost of €6.29–9.38 (base case €8). Similarly, Shen et al. (2023)^44^ applied a micro-costing framework for AI-assisted liquid-based cytology in China, disaggregating consumables, devices, labor, and infrastructure; unit costs declined with scale, from US$10.7 per test at 10,000 women screened to US$5.1 at 100,000. In another study, Wang et al. (2024)^51^ adopted a detailed micro-costing approach within a Markov decision-analytic model to evaluate AI-assisted diabetic retinopathy screening. The total AI screening cost (US$10.65 per person) included recruitment, personnel wages, fundus cameras, engineering and software maintenance, and transportation. AI-specific engineering costs were estimated at US$0.214 per participant, highlighting that algorithm-related expenses were marginal relative to workforce and equipment costs.

Tufail et al. (2016, 2017)^47 48^ conducted a program-level micro-costing analysis of automated retinal image grading within the UK National Health Service (NHS) Diabetic Eye Screening Programme, estimating a per-screening cost of £3.72 for automated grading compared with £4.54 for manual grading. The analysis incorporated software licensing, grading equipment, and staff time, while modeling downstream ophthalmology visits and treatments separately. A similar study by Vargas-Palacios et al. (2023)^50^ performed a detailed bottom-up costing of AI as a second reader in the NHS Breast Screening Programme. AI costs were derived from vendor-quoted fixed and per-screen licensing fees (£35,000 per site per year plus £4.72 per screen), together with infrastructure, training, and maintenance costs annualized over a five-year lifecycle. The estimated mean AI cost was £5.10 per screening, decreasing to around £3.00 at higher volumes, reflecting economies of scale. Earlier work by Guerriero et al. (2011)^19^ similarly applied micro-costing within a UK breast cancer screening trial, capturing radiologist and radiographer time, film and image costs, equipment amortization, and administrative overheads, while also accounting for cost offsets from reduced double reading. Additional examples of micro-costing include Ahmed et al. (2025)^30^, Hu et al. (2024)^35^, Li et al. (2023)^65^, Vargas-Palacios et al. (2023)^50^, Wang et al. (2024)^51^, and Xie et al. (2020)^62^, which incorporated explicit vendor-quoted software licensing, infrastructure, or training costs into model-based frameworks.

Lifecycle costing—explicitly accounting for development, deployment, and maintenance—was identified in only four studies (n = 4)^16 39 51 65^. For example, Vargas-Palacios et al. (2023)^50^ annualized set-up, IT integration, training, and vendor support over a five-year horizon in the NHS Breast Screening Program, whereas van Leeuwen et al. (2021)^49^ modelled only a per-use software fee (US$40 per analysis) and excluded development or capital set-up. Likewise, Schwendicke et al. (2022)^66^ itemized development and deployment but did not include decommissioning or replacement, underscoring how rarely full lifecycle accounting is undertaken.

The cost components captured were heterogeneous. Development and data-generation costs were explicitly itemized in Schwendicke et al. (2022)^66^; clinical validation and regulatory preparation in the same; systems integration and IT infrastructure in Vargas-Palacios et al. (2023)^50^; personnel training in Vargas-Palacios et al. (2023)^50^; and hardware/software expenditures in Fuller et al. (2022)^18^ and Tufail et al. (2016/2017)^47 48^. Maintenance or vendor support appeared intermittently (e.g., annual support in Vargas-Palacios^50^), while periodic model recalibration or retraining was typically omitted (e.g., Fuller 2022^18^). Electricity, cloud compute, and data-hosting costs were rarely specified (e.g., van Leeuwen 2021^49^; Tseng 2021^46^ assumed no incremental AI operating cost). End-of-life costs were almost never addressed; neither Tufail et al.^47 48^ nor Vargas-Palacios et al.^50^ explicitly costed decommissioning or replacement beyond the assumed asset life.

### Pricing Strategies and Reporting

Reporting of pricing strategies was highly variable, with many studies providing only limited or incomplete information (Table 3). Among the included articles, 11 explicitly reported pricing details, while the remainder either omitted such information or presented only general cost summaries without specifying the basis of price.

**Table 3:**
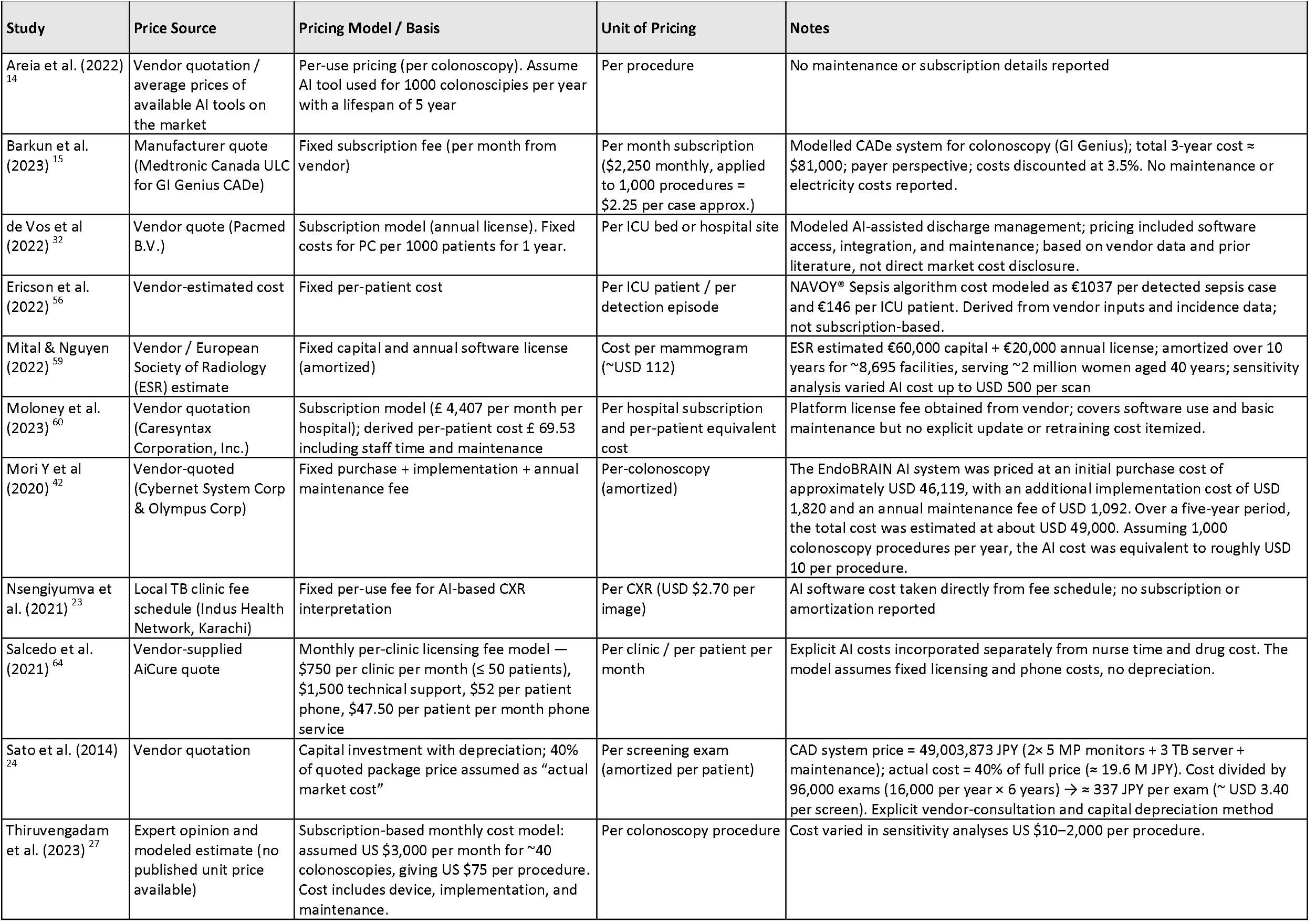
Pricing Strategies and Reporting.

| Study | Price Source | Pricing Model / Basis | Unit of Pricing | Notes |
| --- | --- | --- | --- | --- |
| Areia et al. (2022)<sup>14</sup> | Vendor quotation / average prices of available AI tools on the market | Per-use pricing (per colonoscopy). Assume AI tool used for 1000 colonoscopies per year with a lifespan of 5 year | Per procedure | No maintenance or subscription details reported |
| Barkun et al. (2023)<sup>15</sup> | Manufacturer quote (Medtronic Canada ULC for GI Genius CAdE) | Fixed subscription fee (per month from vendor) | Per month subscription (\$2,250 monthly, applied to 1,000 procedures = \$2.25 per case approx.) | Modelled CAdE system for colonoscopy (GI Genius); total 3-year cost ≈ \$81,000; payer perspective; costs discounted at 3.5%. No maintenance or electricity costs reported. |
| de Vos et al (2022)<sup>32</sup> | Vendor quote (Pacmed B.V.) | Subscription model (annual license). Fixed costs for PC per 1000 patients for 1 year. | Per ICU bed or hospital site | Modeled AI-assisted discharge management; pricing included software access, integration, and maintenance; based on vendor data and prior literature, not direct market cost disclosure. |
| Ericson et al. (2022)<sup>56</sup> | Vendor-estimated cost | Fixed per-patient cost | Per ICU patient / per detection episode | NAVOY® Sepsis algorithm cost modeled as €1037 per detected sepsis case and €146 per ICU patient. Derived from vendor inputs and incidence data; not subscription-based. |
| Mital & Nguyen (2022)<sup>59</sup> | Vendor / European Society of Radiology (ESR) estimate | Fixed capital and annual software license (amortized) | Cost per mammogram (~USD 112) | ESR estimated €60,000 capital + €20,000 annual license; amortized over 10 years for ~8,695 facilities, serving ~2 million women aged 40 years; sensitivity analysis varied AI cost up to USD 500 per scan |
| Moloney et al. (2023)<sup>60</sup> | Vendor quotation (Caresyntax Corporation, Inc.) | Subscription model (£ 4,407 per month per hospital); derived per-patient cost £ 69.53 including staff time and maintenance | Per hospital subscription and per-patient equivalent cost | Platform license fee obtained from vendor; covers software use and basic maintenance but no explicit update or retraining cost itemized. |
| Mori Y et al (2020)<sup>42</sup> | Vendor-quoted (Cybernet System Corp & Olympus Corp) | Fixed purchase + implementation + annual maintenance fee | Per-colonoscopy (amortized) | The EndoBRAIN AI system was priced at an initial purchase cost of approximately USD 46,119, with an additional implementation cost of USD 1,820 and an annual maintenance fee of USD 1,092. Over a five-year period, the total cost was estimated at about USD 49,000. Assuming 1,000 colonoscopy procedures per year, the AI cost was equivalent to roughly USD 10 per procedure. |
| Nsengiyumva et al. (2021)<sup>23</sup> | Local TB clinic fee schedule (Indus Health Network, Karachi) | Fixed per-use fee for AI-based CXR interpretation | Per CXR (USD \$2.70 per image) | AI software cost taken directly from fee schedule; no subscription or amortization reported |
| Salcedo et al. (2021)<sup>64</sup> | Vendor-supplied AiCure quote | Monthly per-clinic licensing fee model — \$750 per clinic per month (≤ 50 patients), \$1,500 technical support, \$52 per patient phone, \$47.50 per patient per month phone service | Per clinic / per patient per month | Explicit AI costs incorporated separately from nurse time and drug cost. The model assumes fixed licensing and phone costs, no depreciation. |
| Sato et al. (2014)<sup>24</sup> | Vendor quotation | Capital investment with depreciation; 40% of quoted package price assumed as “actual market cost” | Per screening exam (amortized per patient) | CAD system price = 49,003,873 JPY (2× 5 MP monitors + 3 TB server + maintenance); actual cost = 40% of full price (≈ 19.6 M JPY). Cost divided by 96,000 exams (16,000 per year × 6 years) → ≈ 337 JPY per exam (≈ USD 3.40 per screen). Explicit vendor-consultation and capital depreciation method |
| Thiruvengadam et al. (2023)<sup>27</sup> | Expert opinion and modeled estimate (no published unit price available) | Subscription-based monthly cost model: assumed US \$3,000 per month for ~40 colonoscopies, giving US \$75 per procedure. Cost includes device, implementation, and maintenance. | Per colonoscopy procedure | Cost varied in sensitivity analyses US \$10–2,000 per procedure. |

Where described, the dominant strategies included subscription-based licensing (n = 6)^15 27 32 42 59 60^, per-use tariffs (n = 2)^14 23^, and institutional or clinic licensing arrangements (n = 2)^56 64^. A smaller number of studies reported prices determined through procurement frameworks or negotiated agreements with public or private payers (n = 1)^24^.

In particular, several recent U.S. studies have adopted subscription-like models for pricing AI systems, reflecting current industry practices. For example, Thiruvengadam et al. (2023)^27^ modeled a monthly subscription cost of USD 3,000 for real-time computer-aided detection (CAD) in colonoscopy, assuming 40 procedures per unit and a derived per-procedure cost of USD 75, which incorporated implementation, device, and maintenance expenses. This approach, derived from expert consultation and market input, typifies the emerging trend toward subscription-based pricing rather than single-use charges. Moloney et al. (2023) described the Caresyntax surgical site infection risk-prediction platform under a fixed subscription model of £4,407 per month per hospital, derived directly from a vendor quote and averaged to £69.53 per patient including staff time in the NHS.

By contrast, Sato et al. (2014)^24^ adopted a capital investment pricing approach for CAD in mammography screening, starting from a quoted package price of 49,003,873 JPY (including two 5-megapixel monitors, a 3-terabyte server, and full maintenance). The authors assumed that only 40% of this full price reflected the “actual market cost,” based on consultation with vendors. They then applied a 6-year lifetime and 50% declining-balance depreciation, spreading the cost over 96,000 screening exams (16,000 women annually), which yielded an amortized per-exam CAD cost of approximately 337 JPY.

### Changes in Clinical Delivery and Cost Offsets

Only a small number of studies explicitly addressed cost offsets or downstream economic effects. Among these, several identified savings arising from reduced hospital admissions, fewer diagnostic tests, or improved clinician efficiency and workflow. Du et al. (2025)^33^, for instance, incorporated offset effects in their model of AI-assisted prostate cancer pathology, capturing savings from reduced diagnostic errors, more appropriate treatment allocation, and diminished pathologist workload. Similarly, de Vos et al. (2022)^32^ modeled AI-supported discharge management in intensive care, where reductions in ICU readmissions and shorter lengths of stay contributed to measurable cost savings.

In ophthalmology, Liu et al. (2023)^41^ showed that AI-enabled telemedicine screening decreased reliance on ophthalmologists and graders, doubling screening throughput (36,000 vs. 15,000 annually) and lowering per-person costs. Similarly, Xiao et al. (2021)^61^ found that AI-assisted glaucoma screening reduced per-person screening costs (US$19.53 vs. US$25.46) primarily through decreased clinician time, although the study did not disaggregate cost components. Xie et al. (2020)^62^ reported workflow-related cost offsets in Singapore’s national diabetic retinopathy screening programme, where automation substantially reduced grader workload and staff requirements. These operational efficiencies translated into lower per-person screening costs (US$62–66 vs. US$77 for human grading), although downstream clinical savings were not modeled.

### Reporting Standards and Cost Estimation Protocols for AI Interventions

We propose a 20-item reporting checklist for costing and pricing of AI-based health interventions (Table 4). The checklist spans five domains—general study information, pricing analysis, costing analysis, cost offsets and savings, and transparency and robustness. It highlights the importance of clearly defining the AI intervention and comparator, stating the analytic perspective and time horizon, and disclosing pricing models and reimbursement arrangements. It also encourages full reporting of lifecycle cost components, including infrastructure, computational, and energy use, and recommends systematic assessment of cost offsets and downstream effects to support comprehensive, policy-relevant evaluations.

**Table 4:** 20-Items Checklist to Guide Costing and Pricing Methodology for AI-Based Health Interventions.

| Domain | Checklist Item | Reported?<br>(Yes/No/Partial) | Page |
| --- | --- | --- | --- |
| General study information | (1) Define the AI intervention (type, function, context, deployment). |  |  |
|  | (2) Specify comparator(s) (usual care or alternative). |  |  |
|  | (3) State perspective (provider, payer, health system, societal) and justify deviations. |  |  |
|  | (4) Define time horizon and discounting. |  |  |
|  | (5) Describe healthcare setting relevant to costing (e.g. volume, staffing, infrastructure). |  |  |
| Pricing analysis | (6) Report price basis (vendor quote, tariff, tender, market). |  |  |
|  | (7) Describe pricing model (per-use, license, subscription, bundled, outcome-based) and tiered/volume elements. |  |  |
|  | (8) Specify pricing duration/unit (annual, monthly, per patient). |  |  |
|  | (9) Indicate if pre- or post-insurance reimbursement/subsidy. |  |  |
|  | (10) State if pricing includes recurrent items (updates, hosting, integration, cybersecurity, support, cloud). |  |  |
| Costing analysis | (11) Report costing approach (micro, gross, ABC, TDABC, hybrid) and rationale. |  |  |
|  | (12) Distinguish average vs incremental; fixed vs variable costs. |  |  |
|  | (13) Provide breakdown across AI lifecycle (development, data prep, validation, regulation, implementation, integration, training, maintenance, recalibration, decommissioning). |  |  |
|  | (14) Describe how unit costs were valued (accounting data, tariffs, market prices, opportunity/shadow costs). |  |  |
|  | (15) State how resource use was measured (observation, system logs, workflow studies, surveys). |  |  |
|  | (16) Report electricity/energy use, hardware depreciation, server or cloud fees. |  |  |
|  | (17) Specify costing reference year and adjustments (inflation, currency conversion, exchange rates if cross-country). |  |  |
| Cost offsets and savings | (18) Report cost offsets (avoided admissions, reduced tests, time saved, error reduction) and how valued. |  |  |
|  | (19) Note unintended financial impacts (induced demand, workflow disruption, staffing changes, spillovers). |  |  |
| Transparency and robustness | (20) Report sensitivity analyses, describe uncertainty, and disclose funding/conflicts of interest (esp. AI vendor ties). |  |  |

We developed the AI Costing Inventory and Calculator (*Supplementary File 2*) as a structured framework to quantify both the monetary costs and the underlying resources use of AI interventions across the technology lifecycle. The tool disaggregates capital investment, development, deployment, and maintenance expenditures into analytically distinct domains, including algorithm design, data infrastructure, clinical validation and regulatory compliance, workforce training, hardware and energy requirements, interoperability and integration layers, platform services, procurement and contracting, and vendor dependencies^9 68-71^. Beyond financial estimates, the inventory now incorporates quantitative indicators of resource use, such as staff hours, personnel skill levels, computational time, and data volume, to reflect the true inputs consumed by each activity. This resource-based approach strengthens the analytical validity and transferability of economic evaluations by anchoring cost estimates in measurable production inputs, thereby distinguishing underlying resource utilization from jurisdiction-specific price differentials.

## DISCUSSION

### Principal findings

This systematic review highlighted a critical methodological shortcoming in the current economic evaluation literature on AI interventions in healthcare. While pricing strategies—such as subscription models, negotiated fees, and benchmarking against comparable products—are frequently reported, the underlying costing approaches remain underdeveloped and inconsistently applied. Most studies provide only limited descriptions of resource use and cost structures, omitting comprehensive accounting of development, implementation, maintenance, and lifecycle costs that are essential for robust and transparent economic evaluation.

Our findings align with the recent systematic review by El Arab et al (2025) ^12^, which concluded that while AI applications often demonstrate favorable incremental cost-effectiveness ratios, methodological weaknesses—particularly incomplete cost reporting, exclusion of infrastructure and indirect costs, and reliance on static models. Similarly, von Gerich et al. (2025)^13^ found that evaluation of machine-learning applications show poor adherence to CHEERS-AI reporting standards, averaging below 40 percent compliance and rarely incorporating the full AI lifecycle of implementation, ongoing maintenance, and model recalibration.

Approximately half of the included studies focused on pricing and half on costing, reflecting a strong dependence on data availability. From an HTA perspective, costing analyses are preferable because they enable greater transparency and reproducibility.

Pricing studies rarely included critical details. Information such as subscription duration, frequency of fees, number of patients or cases covered, and the scale of service delivery was often omitted. For pricing evidence to be policy-relevant, studies should specify subscription arrangements and clearly describe unit costs and service volumes.

Costing studies showed similar limitations. Existing costing studies often did not capture the full range of relevant components, overlooking development costs, integration and implementation expenditures, training requirements, and ongoing maintenance or recalibration. Lifecycle costing, which would account for recurring updates, monitoring, and eventual obsolescence, was largely absent.

Our findings indicate that only a small number of evaluations considered workforce substitution or reallocation, despite the potential for AI systems to shift clinical responsibilities and generate productivity gains or losses. Similarly, long-term health benefits and downstream cost offsets were seldom modelled, with most studies focusing narrowly on immediate or short-term costs. A recurring theme across the literature is the treatment of AI as a one-time investment rather than a continuous service. Only few studies acknowledged the need for recurrent maintenance, periodic updates, and ongoing operational support—factors that are fundamental to sustaining model performance in clinical practice. Moreover, none of the included studies accounted for the substantial energy demands of AI systems, such as electricity consumption, despite the growing importance of these costs from both economic and environmental perspectives.

### Strength and Future Research

To our knowledge, this is one of first systematic reviews to synthesize how costs and pricing of AI interventions in healthcare have been reported, while proposing a structured reporting checklist and a lifecycle-based costing inventory to guide future evaluations. A key strength lies in the comprehensive scope of evidence, covering multiple clinical domains, analytic perspectives, and AI modalities, while simultaneously identifying methodological gaps that undermine transparency and comparability. By distinguishing between costing and pricing, and by itemizing lifecycle components across development, implementation, and maintenance, the review offers a practical framework that complements established HTA standards such as CHEERS 2022^72^ and CHEERS-AI 2024^10^.

### Limitation

Looking forward, several areas warrant priority attention. First, future evaluations should stratify by the type of AI methodology (e.g., shallow vs. deep learning), since computational complexity and retraining requirements imply distinct cost structures and pricing strategies. Second, greater attention is needed to incorporate probabilistic cost-effectiveness analysis, workforce reallocation, and system-level impacts, including equity, access, and environmental externalities such as energy use. Third, the proposed checklist should be piloted in prospective HTA submissions and field-tested across different healthcare settings to refine its feasibility, alignment, and comparability. Lastly, cross-country benchmarking will be essential to elucidate how procurement frameworks, labor costs, and reimbursement systems influence the affordability scalability, and long-term sustainability of AI interventions.

Additionally, we observed considerable inconsistency in the choice of discount rates and time horizons, parameters that critically influence cost-effectiveness estimates. Such variation undermines comparability across studies and may bias the assessment of interventions intended for long-term or global deployment.

A further methodological consideration concerns whether cost-effectiveness analyses of AI interventions adopt deterministic or probabilistic approaches. Most of the studies included in this review relied on deterministic sensitivity analyses, varying single parameters or limited ranges to test robustness, but rarely incorporated full probabilistic cost-effectiveness analysis that captures joint parameter uncertainty across costs, outcomes, and clinical effects. Our findings further support the argument that accurate costing of AI interventions cannot be confined to the algorithm itself. Successful translation of AI into clinical practice depends not only on model performance but also on factors beyond the algorithm — including data lifecycles, deployment infrastructure, end-user integration, and ongoing impact evaluation^73^. Costing frameworks that fail to account for these components risk underestimating the true resource requirements and economic implications of AI implementation In developing our checklist, we placed primarily emphasis on the transparency and completeness of cost component reporting, without explicitly require probabilistic modelling and sensitivity analysis.

A key limitation of our costing inventory is that it primarily reflects early-stage or laboratory-based resource use, whereas real-world deployment of AI systems often occurs within complex commercial and infrastructural ecosystems. In practice, AI is rarely adopted as a standalone algorithm but as part of an “AI-as-a-service” model that requires substantial capital investment in interoperable infrastructure, enterprise integration layers, cybersecurity, and data governance frameworks. These structural requirements represent significant entry barriers and impose recurrent costs that extend well beyond algorithm development, licensing, or maintenance. Importantly, some of these resource requirements, particularly those associated with deployment infrastructure, interoperability, and platform services, may be substantially reduced if governments or health systems invest in shared digital health platforms and enabling ecosystems that lower barriers to scalable AI implementation.

## Conclusion

Our review underscores that many current evaluations of AI interventions still neglect essential cost dimensions, from lifecycle expenditures and incremental costs to downstream cost offsets, thereby limiting their validity and policy relevance. From a policy perspective, investment in shared digital health infrastructure and enabling ecosystems can substantially reduce duplication, lower barriers to deployment, and create the conditions for scalable AI integration. From a research perspective, future studies should adopt more comprehensive costing frameworks, explicitly account for ongoing maintenance and energy demands, and incorporate probabilistic approaches to capture uncertainty.

## Supporting information

Costing Inventory Calculator

## Data Availability

All data produced in the present work are contained in the manuscript.

## Data availability

All data analysed in this review were derived from the published studies cited in the References. The complete data extraction table is provided in the Supplementary Material.

## Code availability

The full search strategies, including all codes and search terms used for database queries, are reported in Supplementary Table 1. Search outputs can be obtained from the corresponding author upon reasonable request.

## Author Contributions

JTL conceived the study, conducted the data analysis, served as second reviewer of the search, and drafted the initial manuscript. TS performed the systematic review search. VL acted as third reviewer of the search and assisted in validating the cost estimation calculator. TS and CWH further contributed to data extraction and literature summarization. All authors reviewed and revised the manuscript critically for important intellectual content and approved the final version.

## Acknowledgements

We acknowledge the contributors to the International Prospective Register of Systematic Reviews (PROSPERO). JTL was partially supported by the Ministry of Education, Taiwan, through the Yushan (Mount Jade Scholar) Fellow Program (MOE-112-YSFMN-0003-002-P1).

## TABLE and FIGURES

**Supplementary Table S1:**
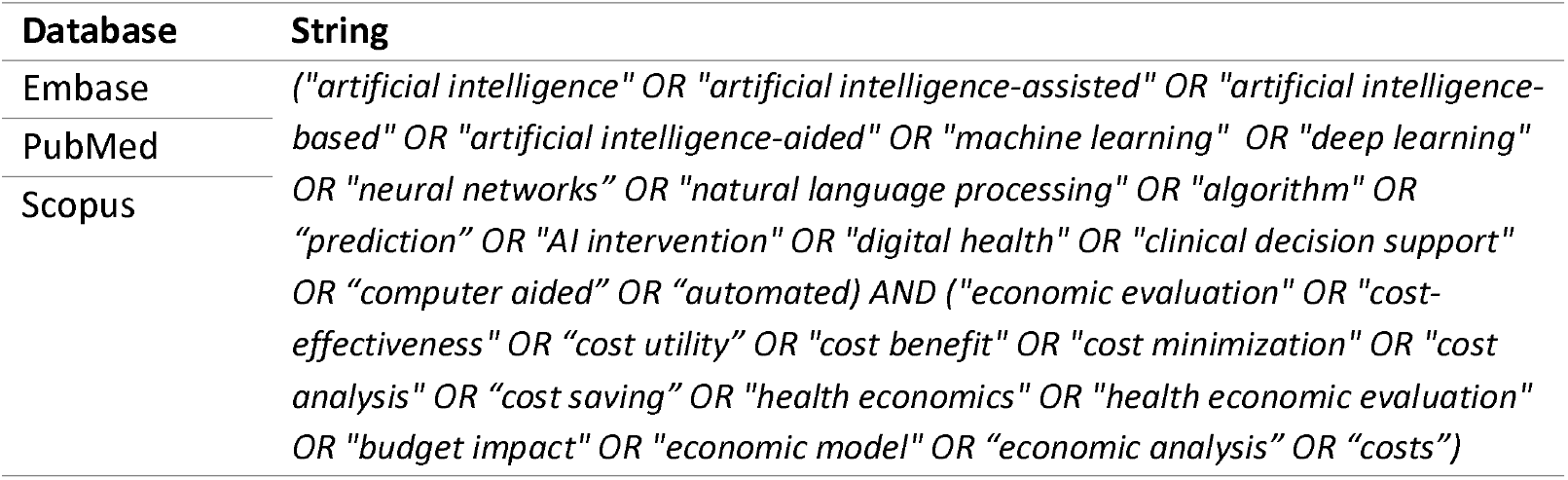
Search Strategies for Economic Evaluation of AI Interventions.

**Supplementary Table S2:** Overview of Costing and Economic Evaluation Studies of AI-Based Interventions.

| Study | Clinical Area / Condition of Interest | Country | Target Population | AI Intervention Assessed | Economic Evaluation Type | Perspectives | AI Costing or Pricing Reported | Key Findings in terms of costs for AI Intervention | Included activities |
| --- | --- | --- | --- | --- | --- | --- | --- | --- | --- |
| Adams et al. (2021)<sup>54</sup> | Lung cancer | USA | Adults undergoing baseline low-dose CT lung cancer screening | AI-based malignancy risk prediction score combined with Lung-RADS for lung nodule classification | Cost-consequence analysis | Healthcare payer | Not reported | N/A |  |
| Areia et al. (2022)<sup>14</sup> | Colorectal cancer | USA | Hypothetical cohort of 100 000 average-risk adults aged 50–100 years | AI-aided colonoscopy screening | Cost-effectiveness analysis | Societal | Pricing | US \$ 19 per procedure, based on the average price of available AI tool on the market | Cost of AI software per colonoscopy; screening and surveillance colonoscopy; polypectomy; pathology; colorectal cancer treatment (stage-specific); adverse events; overhead |
| Ahmed et al. (2025)<sup>30</sup> | Pediatric diabetic retinal disease (DRD) | USA | Under aged 21 years pediatric patients with Type 1 and Type 2 diabetes | Autonomous AI system for DRD screening | Cost-effectiveness analysis | Healthcare provider | Costing | Cost of the A-assisted DRDI screening varied by health system size: \$242 additional per patient to -\$140 savings; follow-up adherence costs \$1,106 to -\$588 savings | Screening (AI acquisition, integration, support, space, operator labor); follow-up (ECP exam costs if within health system) |
| Barkun et al. (2023)<sup>15</sup> | Colorectal adenoma/CRC screening | Canada | Patients ≥50 years undergoing colonoscopy following a positive fecal immunochemical test | AI-aided colonoscopy (GI Genius CAdE) versus conventional colonoscopy | Cost-utility Analysis | Healthcare payer | Pricing | cost of the CAdE solution was estimated at \$2250 for a monthly subscription | Aggregate direct medical costs (colonoscopy, FIT, polypectomy, surgery, chemotherapy, follow-up visits) |
| Bashir et al. (2022)<sup>16</sup> | Pulmonary tuberculosis | Pakistan | Individuals screened for TB via chest X-ray | CAD software for CXR interpretation | Costing analysis | Healthcare payer | Costing | Per-screen CAD costs varied by software and throughput: CAD4TB (0.25–2.33 USD) and InferRead | Equipment (hardware, licenses, data storage), installation, support/maintenance |
|  |  |  |  |  |  |  |  | (0.19–2.78 USD) were cheaper than radiologists (0.70–0.93 USD) at higher volumes; Lunit (0.94–1.69 USD) and qXR (0.95–1.90 USD) only competitive at very high volumes. | e; Radiologist time, training; software training |
| Bustamante-Balen et al. (2025)<sup>31</sup> | Colorectal cancer screening/ Adenoma detection | Spain | Adult patients eligible for colonoscopy | GI Genius Intelligent Endoscopy Module for CAdE/CAD | Cost-effectiveness analysis | Healthcare payer | Not clear | Assumption of € 7.59/per colonoscopy | N/A |
| Curl et al. (2023)<sup>17</sup> | Osteoporotic vertebral compression fractures (OVCFs) | USA | Adults (base case: 71-year-old woman with chest/abdominal radiographs, no prior osteoporosis diagnosis) | AI-based deep learning software for opportunistic fracture screening on existing radiographs | Cost-utility analysis | Societal | Not clear | Base-case AI screening cost assumed \$10 per patient, varied in sensitivity analysis from \$1–100 | Software implantation cost per patient screened (\$10 in base case) and literature-derived estimates of fracture treatment and long-term care costs (all adjusted to 2022 USD) |
| de Vos et al (2022)<sup>32</sup> | Intensive care / ICU | Netherlands | Adult ICU patients; model focuses on those eligible for ICU discharge | Pacmed Critical (ML decision-support predicting 7-day combined risk of ICU readmission or mortality to guide discharge timing) | Cost-utility analysis | Societal | Pricing | Included a one-off acquisition price for PC (modeled per 1,000 patients/year) in base case | Unit costs (e.g., per ICU- and ward-day, procedure) obtained from Amsterdam UMC accounting data, supplemented by literature and expert opinion |
| Deladillo et al (2021)<sup>55</sup> | Mental health / depression and anxiety disorders | UK | Adults receiving stepped-care psychological therapy | Machine learning–based stratified care model | Cost-effectiveness analysis | Healthcare payer | Not reported | N/A |  |
| Du et al. | Prostate | Sweden | Men aged 50–74 | AI-assisted pathology | Cost-utility | Healthcare | Not clear | The base-case AI | Applied aggregated |
| (2025)<sup>33</sup> | cancer |  | undergoing quadrennial PSA screening |  | analysis | payer |  | cost was assumed to be €10 per patient (case). | per-case unit costs (e.g., €10 per AI-assisted case; standard pathology cost) drawn from Swedish health-care pricing data |
| Ericson et al. (2022)<sup>56</sup> | Sepsis (early detection in ICU) | Sweden | Adult ICU patients not diagnosed with sepsis at admission | Machine-learning sepsis prediction algorithm (InSight™) | Cost-effectiveness analysis | Healthcare payer | Pricing | The annual licensing fee of sepsis prediction algorithm is €1,037 per bed. | Costs of the prediction algorithm; ICU and general-ward days; readmissions; long-term consequences (impaired kidney function, amputation, depression, PTSD) |
| Fuller et al (2022)<sup>18</sup> | Diabetic Retinopathy | USA | Low-income adults with diabetes receiving primary care | Automated Retinal Image Analysis System (ARIAS) for DR screening | Cost-utility analysis | Healthcare payer | Costing | US\$31.02 per screened patient (includes camera, maintenance, software, labor, etc.) | |
| Gomez Rossi et al. (2022)<sup>34</sup> | Melanoma, Dental Caries, Diabetic Retinopathy | USA, Germany, Brazil | US adults aged 50 (melanoma), German children aged 12 (dental caries), Brazilian adults aged 40 with diabetes (retinopathy) | AI-based screening and diagnostic support | Cost-effectiveness analysis | Societal perspective | Costing | For the dental use case, an €8 fee per application had been assumed in the original publication based on direct costs for research | Aggregate per-patient (dermatology/ophth almolgy) or per-tooth (dentistry) cost estimates for detection, diagnostic confirmation, treatment, and follow-up. AI was modelled as a fee-for-service (€8/unit), with all costs converted via 2020 PPP. |
| Guerriero et al. (2011) <sup>19</sup> | Breast cancer screening (mammography) | UK | Women aged 50–70 undergoing routine NHS breast screening | Single reading with Computer-Aided Detection (CAD) | Cost-effectiveness analysis | Healthcare payer | Costing | Incremental cost (compared with double reading by radiologists) per woman screened ranged from £227–£590 depending on center size, including savings from reduced radiologist reading time | CAD equipment purchase, upgrade and maintenance; reader training; reading time (cost/minute × time); recall-assessment costs (imaging, biopsy) |
| Hill et al. (2020) <sup>57</sup> | Atrial Fibrillation (screening and stroke prevention) | UK | Adults ≥50 years, eligible for AF screening | Machine learning risk prediction algorithm to identify high-risk patients for targeted screening | Cost-effectiveness analysis | Healthcare payer | Not reported | N/A |  |
| Hu et al. (2024) <sup>35</sup> | Diabetic retinopathy screening | Australia | Adults with diabetes aged ≥20 years | AI-based diabetic retinopathy screening (Including 3 scenarios: replace manual grading; scale-up to patient acceptance; universal screening) | Cost-effectiveness analysis | Societal | Costing | AI intervention costs estimated AU\$12.20 per screening for AI-based DR screening | Screening costs (AI license, hardware depreciation, operator labor), link-to-care costs, direct medical costs (consultations, angiography, OCT, photocoagulation, anti-VEGF injections, vitrectomy), and blindness care costs |
| Huang et al. (2025) <sup>36</sup> | Lung cancer screening | China (Fujian Province, rural underserved areas) | Socioeconomically marginalized adults ≥40 years who had not previously undergone LDCT screening | Mobile low-dose CT screening with an AI-assisted diagnostic system | Cost-effectiveness analysis | Societal | Not reported | N/A | Total program costs (¥1,203,504 / US\$188,847.71) were summed and divided by the number of screenings to derive an average cost per screening (¥118.47 / US\$18.65). Cost-effectiveness ratios (CERs) were then calculated as cost per suspected (¥11,143.56 / US\$1,753.29) and confirmed lung cancer detected (¥16,950.76 / US\$2,669.94). |
| Huang et al. (2022)<sup>20</sup> | Diabetic retinopathy | China | Newly diagnosed diabetes patients without DR (mean age 44) | AI-based diabetic retinopathy screening | Cost-effectiveness analysis | Healthcare provider/ Societal | Not clear | The AI software is estimated at \$ 1.45 per screening | Screening (AI software fees; salaries of eye-care staff; equipment maintenance; advertising; building rent), follow-up exams (fundus imaging, visual acuity, intraocular pressure, slit-lamp), laser treatment; direct non-medical (transport); indirect (patient and caregiver productivity losses; blindness costs) |
| Kacew et al (2021)<sup>37</sup> | Colorectal cancer | USA | Hypothetical cohort of patients with newly diagnosed de novo metastatic CRC | Digital histopathology AI algorithm to determine mismatch repair (MMR) / microsatellite instability (MSI) status | Cost-effectiveness analysis | Healthcare payer | Not clear | AI image scanning cost was US\$6.07 per patient | |
| Li et al (2023)<sup>38</sup> | Diabetic Retinopathy (DR) screening | China (rural Shanxi) | Adults with type 1 or type 2 diabetes (hypothetical cohort of 10,000 patients, aged ≥18 | AI-based DR screening (EyeWisdom deep learning system) | Cost-utility analysis | Healthcare payer | Costing | Fundus camera (USD 27,234, 8-year lifespan) and AI software (USD 13,350, 50-year lifespan). |  |
| Libanio et al | Gastric | Europea | Average-risk adults | AI-assisted | Cost-utility | Societal | Not | N/A |  |
| (2023) <sup>21</sup> | cancer and colorectal cancer | n countries (Portugal, Italy, Netherlands) | undergoing screening endoscopy/colonoscopy | esophagogastroduodenoscopy (EGD) for gastric cancer detection, integrated with colorectal cancer colonoscopy screening | analysis |  | reported |  |  |
| Lin et al (2024) <sup>39</sup> | Fundus diseases screening | China (Shanghai) | Older population (community residents aged ≥60; modeled cohort of 100,000) | AI-assisted fundus disease screening (telemedicine + AI interpretation vs. manual grading) | Cost-utility analysis | Societal | Costing | Screening costs included labor, equipment, and software, with equipment and software estimated at \$1.40 per screening. | |
| Lin et al (2023) <sup>40</sup> | Diabetic Retinopathy (DR) screening | China (Shanghai) | Older adults with diabetes (modeled cohort age 65+, community-based residents) | AI-assisted DR telemedicine screening | Cost-utility analysis | Societal | Costing | The cost difference between AI-assisted and manual screening was USD 0.50 per person. |  |
| Liu et al. (2023) <sup>41</sup> | Multiple blindness-causing eye diseases (age-related macular degeneration, glaucoma, diabetic retinopathy, cataract, pathological myopia) | China | Adults aged ≥50 (hypothetical cohort, modeled for rural and urban settings) | AI-based telemedicine screening (2) non-AI telemedicine screening; (3) AI telemedicine screening | Cost-utility analysis | Societal | Costing | Annualized AI telescreening cost: US\$6 (rural) and US\$8 (urban) per person. Detailed costing included: staffing assumptions (eg, 6 staff for non-telemedicine, 2 staff for AI telescreening), equipment costs (tonometer US\$11,958; auto-refractometer US\$20,000; retinal camera US\$27,035; slit lamp US\$1,007; chart US\$278; laptops US\$2,552), telemedicine/AI platform development and maintenance, and throughput differences (AI 36,000 vs. manual 15–24,000 patients/year). | Screening [face-to-face, non-AI telemedicine, AI telemedicine], definitive ophthalmic check, intervention [consultation, medications, surgery], follow-up visits), direct non-medical costs (transport, food, accommodation for patients/family), and indirect costs (patient and caregiver productivity losses) |
| Liu et al (2024) <sup>58</sup> | Asymptomatic left ventricular dysfunction (LVD, LVEF ≤40%)<br>Asymptomatic Left Ventricular Dysfunction | Taiwan | Adults (≥18 years, focus on age 65 in cost-effectiveness base case) undergoing ECG at outpatient clinics or health check-ups | AI-enabled electrocardiography (AI-ECG) algorithm for opportunistic screening and risk stratification of asymptomatic LVD | Cost-effectiveness analysis | Healthcare provider | Not reported | AI-ECG screening cost was assumed equivalent to a standard ECG: US\$4.96 (base case), with sensitivity analysis up to 500% higher | |
| Maas et al. (2025) <sup>53</sup> | Hepatocellular carcinoma (HCC) | Italy | Cirrhotic patients at risk of HCC ≥ aged 50 | AI-assisted MRI tool for HCC lesion detection | Cost-effectiveness analysis | Healthcare provider | Not clear | Cost of the AI-assisted MRI tool was estimated at €300 per patient | N/A |
| MacPherson et al (2021) <sup>22</sup> | Diabetic retinopathy | UK (Scotland) | Adults with diabetes invited to the national diabetic eye screening program | AI-based autonomous image grading system for diabetic retinopathy screening compared with manual human grading | Cost-effectiveness analysis | Healthcare payer | Not reported | N/A |  |
| Mervin et al. (2018) <sup>63</sup> | Mental Health / Dementia | Australia | Older adults (≥60 years) with dementia living in long-term care facilities | PARO – therapeutic robotic seal (with artificial intelligence enabled), compared to PARO with AI disabled (plush toy) and usual care | Cost-effectiveness analysis | Healthcare provider | Costing | Equipment: robotic seal (AUD\$23,280), battery (AUD\$1,810), accessories (AUD\$220). Maintenance: robotic maintenance (AUD\$1,445), cleaning (AUD\$443) | Cost of PARO device rental/maintenance, staff time for 15-minute sessions (3×/week for 10 weeks), comparison plush-toy cost, and usual-care resource use |
| Mital & | Breast | USA | Women aged 40– | AI-guided mammography | Cost-Utility | Healthcare | Pricing | Fixed costs of | Costs of risk |
| Nguyen (2022) <sup>59</sup> | cancer screening | | 49 years | screening (risk stratification by deep-learning reading) versus PRS-guided, family history-guided, annual screening for all, and no screening | analysis | provider | | €60,000 (\$65,300) in addition to an annual cost of €20,000 (\$21,770) for the software license. Annual screening for all 2,022,120 (table2) | prediction (AI reading of index mammogram; OncoArray genetic test + pre/post genetic counselling at \$44/session), digital mammography (CMS physician fee schedule), diagnostic work-up after positive screens, and stage-specific breast cancer treatment; AI software license amortized over 10 years (~\$112 per mammogram) |
| Moloney et al. (2023) <sup>60</sup> | Surgical site infection (SSI) undergoing colorectal surgery | UK | Patients undergoing colorectal surgery | AI/machine learning-based predictive algorithm | Cost-utility analysis | Healthcare provider | Pricing | The monthly subscription price of the Caresyntax platform of £4407.00 was obtained from the Caresyntax Corporation | Unit costs for NPWT kit (and replacement), staff (nurse) time, platform subscription (allocated per patient), extended hospital stay (per-day cost), readmission episode cost, antibiotic cost |
| Mori Y et al (2020) <sup>42</sup> | Colorectal polyps / Colorectal cancer prevention | Japan, England, Norway, USA | Patients with diminutive (≤5 mm) rectosigmoid polyps undergoing colonoscopy | AI-assisted optical diagnosis (EndoBRAIN) enabling a “diagnose-and-leave” strategy vs standard resect-all-polyps | Costing analysis | Healthcare payer | Pricing | in Japan only. Initial purchase US\$46,119, implementation US\$1,820, annual maintenance US\$1,092 (total US\$49,031). | Calculated average colonoscopy cost (including pathology) under two management scenarios (diagnose-and-leave |
| | | | | | | | | Assumed 1000 cases/year over 5 years resulting in approximately US\$10 per colonoscopy | vs resect-all) using country-specific public-insurance reimbursement rates; then multiplied by annual colonoscopy volumes to estimate gross annual reimbursement |
| Morrison et al. (2022)<sup>43</sup> | Retinopathy of prematurity (ROP) | USA | Preterm infants | AI-based ROP screening (assistive and autonomous) compared with telemedicine and ophthalmoscopy | Cost-effectiveness analysis | Healthcare payer | Costing | A hypothetical price of \$30 perscreen, which include professional fees and technical fees. | Unit costs derived from Current Procedural Terminology (CPT) codes (professional + technical fees) and estimated physician opportunity costs |
| Nsengiyumva et al. (2021)<sup>23</sup> | Tuberculosis diagnosis | Pakistan | Adults ≥15 years with TB symptoms | AI-based CXR interpretation | Cost-effectiveness analysis | Healthcare provider | Pricing | AI-based CXR interpretation \$2.70 Table3 | Applied programme-level unit costs (2020 US\$) for each diagnostic test (smear, GeneXpert, CXR) and TB treatment—sourced from the Pakistan TB Program—and multiplied by the number of persons simulated |
| Salcedo et al. (2021)<sup>64</sup> | Active (pulmonary) tuberculosis | USA (Los Angeles County) | Adults with non-drug-resistant pulmonary TB | AiCure AI monitoring platform (mobile-based, computer vision) | Cost-effectiveness analysis | Societal | Pricing | Device (\$52/patient), phone service (\$47.5/month), licensing (\$750/50 patients/month per clinic), tech support (\$1,500/month per clinic) | Bottom-up enumeration of resource use: drug acquisition (\$174.16/patient-month); AiCure licensing (\$750/clinic/month), technical support (\$1,500/clinic/month); phone hardware (\$52/patient fixed) and service (\$47.50/patient-month); LVN/RN time (hourly wage plus travel costs) |
| Sato et al. (2014)<sup>24</sup> | Breast cancer screening (mammography) | Japan | Hypothetical cohort of women aged 50, population-based screening | CAD-assisted single reading vs. double reading | Cost-effectiveness analysis | Societal | Pricing | CAD installation cost approximately 337 JPY per exam | Direct medical costs for screening and treatment were applied per hypothetical 50-year-old examinee, rather than detailed bottom-up resource measurement. |
| Schwendicke et al (2021) (2022)<sup>66 67</sup> | Dental caries detection | Germany | Posterior permanent teeth in 12-year-olds | AI-based radiographic detection | Cost-effectiveness analysis | Healthcare provider | Costing | AI cost: €8 per use (sensitivity €4–12) |  |
| Sekiguchi et al. (2023)<sup>25</sup> | Colorectal cancer screening (colonoscopy after FIT) | Japan | Adults 40–74 yrs, average-risk | Colonoscopy with computer-aided detection (CADE) | Cost-effectiveness analysis | Healthcare payer | Not reported | N/A | Costs were estimated using a population-based simulation model that incorporated aggregated (reimbursement-based) cost inputs—including the per-procedure CADE cost |
| Shen M et al. (2023)<sup>44</sup> | Cervical cancer screening | China | Women aged 30–64 | AI-assisted liquid-based cytology screening (Landing CytoScanner) | Cost-effectiveness analysis | Healthcare provider | Costing | US\$5.1/test (100,000 women) | Detailed unit costs for each screening test (manual LBC, HPV-DNA, AI-LBC at varying volumes), colposcopy, biopsy, loop electrosurgical excision procedure (LEEP), hysterectomy, administration, and stage-specific cervical cancer treatment (initial hospitalization + annual follow-up) |
| Srisubat et al. (2023)<sup>26</sup> | Diabetic retinopathy screening | Thailand | Adults with diabetes in the national screening program | Deep-learning automated retinal image grading vs. trained human graders | Cost-utility analysis | Healthcare payer | Not clear | AI grading assumed 32 THB (~US\$1) per patient | Direct medical costs (fundus camera, CFP capture, image interpretation, confirmatory grading, outpatient visits, treatments for sight-threatening DR and DME including bevacizumab injections, laser photocoagulation, vitrectomy); direct non-medical costs (patient food, transport, accommodation, equipment, caregiver productivity loss based on GNI per capita); cost of blindness (visual rehabilitation) |
| Sun MY et al. (2024)<sup>45</sup> | Malnutrition and nutritional diagnosis in | China | Adult inpatients (≥18 years) in 11 hospitals across 10 provinces | AI-based rapid nutritional diagnostic system integrated into routine inpatient care | Cost-effectiveness analysis | Healthcare provider | Not clear | AI assessment cost estimated at 8 CNY vs. manual diagnosis 24.5 CNY | Per inpatient diagnostic, treatment, and care resources |
|  | hospitalised patients |  |  | AI-based rapid nutritional diagnostic system for routine inpatient nutritional assessment and diagnosis |  |  |  |  |  |
| Thiruvengadam et al. (2023)<sup>27</sup> | Gastroenterology – Colorectal cancer (CRC) screening via colonoscopy and adenoma detection | USA | Average-risk adults aged 45 years and older undergoing CRC screening | Real-time Computer-Aided Detection (CAD) system for colonic neoplasia | Cost-effectiveness Analysis | Healthcare provider | Pricing | CAD cost range \$10 – \$2,500 | Literature-derived unit costs (2020 USD) for colonoscopy (screening, biopsy, snare polypectomy), CAD (\$75/procedure), complication management (bleeding, perforation), and colorectal cancer (CRC) care (initial, continuing, terminal) drawn primarily from Medicare fee schedules and published cost studies |
| Tseng et al. (2021)<sup>46</sup> | Asymptomatic Left Ventricular Dysfunction (ALVD) | USA | Adults aged 55–75 years (base case: 65 years), asymptomatic individuals at average risk | Deep learning–based electrocardiographic (AI-ECG) screening algorithm for early detection of asymptomatic LV dysfunction | Cost-effectiveness analysis | Healthcare Payer | Not reported | AI-ECG screening modeled at US\$50 per test, consistent with the Medicare reimbursement rate for standard ECG | Costs included the AI-ECG screening (incremental cost added to a standard ECG), transthoracic echocardiography (TTE), downstream confirmatory tests, heart failure event costs, and long-term treatment costs |
| Tufail et al. (2016) (2017) | Ophthalmology / Diabetic | UK | Adults with diabetes eligible for | Automated retinal image analysis systems (EyeArt, | Cost-effectiveness | Healthcare payer | Costing | The cost of automated screening | Fixed and variable screening costs |
| <sup>47,48</sup> | Retinopathy Screening |  | national screening | Retmarker, iGradingM) used either to replace initial (level-1) manual grading or to pre-screen images before human grading | Analysis |  |  | was estimated at £3.72 per patient compared with £4.54 per patient for manual grading, based on the NHS Diabetic Eye Screening Programme cost structure. Costs included equipment, software licensing, and staff time; downstream ophthalmology visits and confirmatory diagnostic costs were modeled separately. | captured via a detailed survey of the local NHS diabetic eye screening centre—specifically the cost of level-1 manual grading, automated software licence fees (EyeArt, Retmarker, iGradingM), equipment overheads, and follow-up/referral costs generated by false positives |
| Leeuwen et al. (2021) <sup>49</sup> | Acute ischemic stroke – large vessel occlusion (LVO) detection using CT angiography | UK | Adults with suspected acute ischemic stroke eligible for endovascular treatment | AI-assisted vessel occlusion detection software | Cost-effectiveness analysis | Societal | Not clear | Per-use license fee modeled at US\$40 per AI analysis, varied between US\$0–200; no inclusion of development or capital setup costs | Applied published average treatment costs for IAT-eligible (\$11,728) and non-IAT patients (\$1,004), short-term (<90 days) costs by modified Rankin Scale (mRS) health state, and annual long-term costs by mRS; AI priced per analysis (\$40); all inflated to 2019 USD and discounted at 4% |
| Vargas-Palacios et al. (2023) <sup>50</sup> | Breast cancer screening | UK | Adult women (≥ 50 years) in the NHS Breast Screening Programme | Mia® AI system (Kheiron Medical Technologies) used as a second reader replacing one human | Cost-effectiveness analysis | Societal | Costing | AI price £4.72 per scan (including maintenance); set-up cost £35 000 per | Costs of radiologist reading (£5.90/scan), AI set-up (£35k/site), |
|  |  |  |  | reader in double-reading mammography |  |  |  | site; compared to radiologist cost £5.90 per scan. | maintenance, AI price/scan (£4.72), arbitration, recall triple-assessment (clinical exam + imaging ± biopsy), stage-specific treatment costs, screening invitations, locum outsourcing |
| Wang et al. (2024) <sup>51</sup> | Diabetic retinopathy screening | China | Adults with diabetes eligible for national screening | AI-assisted fundus image screening system for automated diabetic retinopathy detection and referral (AI-based DR screening model) | Cost-effectiveness Analysis | Societal | Costing | Total estimated AI-based screening cost: US\$10.65 per person screened | Screening (advertising, imaging equipment, personnel time, AI deployment), referral examination (clinic visits, diagnostic tests), DR treatment (laser, anti-VEGF), transportation and food (patient and companion), income loss (patient and family), blindness care (direct medical, non-medical, indirect) |
| Wolf et al (2020) <sup>52</sup> | Diabetic retinopathy screening | USA | Children / Adolescents (<21 years) with Type 1 or Type 2 Diabetes | FDA-approved Autonomous AI Diagnostic System for Diabetic Retinopathy (Point-of-Care system integrated in pediatric endocrinology clinic) | Cost-effectiveness analysis | Patient / Family perspective (out-of-pocket cost) | Not reported | N/A |  |
| Xiao et al. (2021) <sup>61</sup> | Glaucoma screening in elderly | China | Older adults (≥65 years) in remote or underserved areas | AI-assisted population-based glaucoma screening using deep learning | Costing analysis | Healthcare provider | Costing | AI-assisted screening costed US\$19.53 per person, not itemized | Annualized capital costs of screening equipment; |
|  | populations |  |  | analysis of fundus images |  |  |  | by component | confirmatory diagnostic costs; treatment costs for laser peripheral iridotomy (PACS/PAC) and trabeculectomy (PACG), including inpatient examinations, consumables, nursing, bed-days, and outpatient medication |
| Xie et al. (2020)<sup>62</sup> | Diabetic retinopathy screening | Singapore | Adults with diabetes participating in the national diabetic retinopathy screening programme | Deep learning system (DLS) for automated or semi-automated grading of retinal fundus images within a teleophthalmology-based screening programme | Cost-minimization analysis | Healthcare provider | Costing | The semi-automated AI model achieved a per-person annual screening cost of US\$62, the fully automated model US\$66, compared with US\$77 for human assessment. | Per-patient total cost inputs (screening costs + medical consultation costs converted to US\$) to estimate average annual cost per patient for each screening model |
| Yonazu et al. (2024)<sup>28</sup> | Early Gastric Cancer | Japan | Adults with moderate to severe atrophic gastritis due to H. pylori infection | AI-based computer-assisted diagnosis (CADx) support system "Tango" for endoscopy | Cost-effectiveness Analysis | Healthcare payer | Costing | Installation cost US\$6,000; annual contract US\$10,000; amortized to US\$45 per endoscopy | • Biopsy + pathology (80 USD)<br>• CADx (Tango) per-procedure marginal cost (45 USD; derived from amortized installation/contract fees)<br>• Treatment costs by cancer stage (Stage I: 7,464 USD; II: 14,402 USD; III: 22,736 USD; IV: 195,742 USD) |
| Ziegelmayr et al. (2022)<sup>29</sup> | Lung cancer / Low-dose CT screening | Germany, using U.S. Medicare cost base | Adults aged 55–80 years at high risk | AI-assisted lung nodule detection and classification using 3D CNN integrated with low-dose CT | Cost-effectiveness Analysis | Healthcare payer | Not clear | AI cost up to USD 68 remained cost-saving | Unit costs for: initial low-dose CT (USD 161); follow-up after false-positive (CT + bronchoscopy + biopsy, USD 2,256); curative resection (USD 36,305); annual palliative care (USD 60,000); plus zero costs for “no BC” or death states |

